# Multivariate patterns of disrupted sleep longitudinally predict affective vulnerability to psychosis in 22q11.2 Deletion Syndrome

**DOI:** 10.1101/2023.01.31.23285240

**Authors:** Natacha Reich, Farnaz Delavari, Maude Schneider, Niveettha Thillainathan, Stephan Eliez, Corrado Sandini

## Abstract

22q11.2 deletion syndrome (22q11DS) contributes dramatically increased genetic risk for psychopathology, and in particular schizophrenia. Sleep disorders, including obstructive sleep apnea (OSA), are also highly prevalent, making 22q11DS a unique model to explore their impact on psychosis vulnerability. Still, the contribution of sleep disturbances to psychosis vulnerability remains unclear.

We characterized the sleep phenotype of 69 individuals with 22q11DS and 38 healthy controls with actigraphy and sleep questionnaires. Psychiatric symptoms were measured concomitantly with the baseline sleep assessment and at longitudinal follow-up, 3.58±0.85 years later. We used a novel multivariate partial-least-square-correlation (PLSC) approach to identify sleep patterns combining objective and subjective variables, which correlated with psychiatric symptoms. We dissected longitudinal pathways linking sleep disturbances to psychosis, using multi-layer-network-analysis.

22q11DS was characterized by a non-restorative sleep pattern, combining increased daytime fatigue despite longer sleep duration. Non-restorative sleep combined with OSA symptoms correlated with both emotional and psychotic symptoms. Moreover, a sleep pattern evocative of OSA predicted longitudinal worsening of positive and negative symptoms, by accentuating the effects of emotional dysregulation. These results suggest that sleep disturbances could significantly increase psychosis risk, along an affective pathway. If confirmed, this suggests that systematic screening of sleep quality could mitigate psychosis vulnerability in 22q11DS.

## 1. Introduction

Sleep difficulties occur in up to 80% of individuals diagnosed with a mental health disorder [1]. Still, given seemingly non-specific clinical associations, sleep difficulties has been mostly considered as a secondary consequences of mental health disturbances [1]. Indeed, most conceptual models have considered different psychiatric disorders to be the expression of a series of discrete underlying “disease” mechanisms [2-4]. However, it has recently become clear that early stages in the emergence of psychiatric disorders are not as specific or discrete as it had been previously imagined [5, 6]. For instance, symptoms of emotional dysregulation, such as anxiety or reduced tolerance to stress, strongly increase vulnerability to psychotic disorders among at-risk individuals, along an “affective pathway” towards psychosis [7, 8]. Psychiatric disorders might hence be best understood as complex systems of reciprocally interacting symptoms [2, 9]. Embracing such clinical complexity has spiked interest in identifying disease mechanisms that might bridge across different forms of psychopathology [9-11].

Sleep disturbances could represent a key factor in such complex transdiagnostic clinical trajectories. Indeed, it is now clear that sleep quality can actively influence mental health [12]. Sleep disruption directly modulates limbic functioning, leading to sub-cortical limbic hyperactivity and pre-frontal hypofunction [13]. As a result, disruption, in particular of REM sleep, determines profound alterations in emotional expression and regulation, accentuating negative affect and stress reactivity, diminishing positive affect, and impairing re-organization of emotional memories [13]. From a developmental perspective, early sleep disturbances predict future development of emotional and behavioral problems such as depression, anxiety and ADHD [14] [15-17]. Sleep disruption is also commonly observed in psychotic disorders [18], being identified as a treatment priority by patients [19] and independently predicting quality of life and overall functioning [20]. Moreover, sleep disturbances might actually play an active role in vulnerability to psychosis. Indeed, sleep disorders such as insomnia and nightmare disorders correlate with the severity of psychotic symptoms [21] and represent early predictors of clinical relapse in schizophrenia [22]. Alterations in sleep continuity, sleep duration and circadian rhythm have been recently observed before the first psychotic episodes [23] and might predict longitudinal worsening of positive symptom in youth at Clinical High-Risk (CHR) for psychosis [24-26]. These findings suggest that sleep disturbances could represent a promising target for early preventive interventions in psychosis. To date however, we still have an insufficient understanding of the clinical pathways linking sleep disruption to psychosis, which has limited translation to clinical practice [18]. In the present work, we attempt to address two main research challenges that might have significantly contributed to this lack of knowledge.

A first major research challenge has consisted in obtaining sufficiently precise sleep phenotyping before the onset of full-blown psychotic disorders [27]. Here, we take advantage of the 22q11.2 Deletion Syndrome (22q11DS), which has emerged as a unique genetic model to study early stages of psychosis [28]. 22q11DS is caused by a 1,5-3-Mb deletion on the long arm of the chromosome 22, occurring in 1 of 2148 live birth [29]. Individuals with 22q11DS are typically diagnosed at a young age, due to a complex phenotype, including congenital heart disease, velopharyngeal insufficiency and learning difficulties [30]. Moreover, the syndrome is associated with a unique psychiatric phenotype, characterized by a 30-40% risk of developing a psychotic disorder by adulthood [31]. Aside from psychosis, various other forms of psychopathology, including ADHD, anxiety and mood disorders are also strongly increased in 22q11DS [31]. As in the general population, emotional dysregulation increases risk of psychosis in 22q11DS [8, 32-35]. To date, few studies have investigated sleep in 22q11DS and its contribution to the clinical phenotype typically observed in this population. However, available evidence indicates that sleep disorders might be over-represented [36]. Subjective complaints of sleep onset delay, agitated sleep, and excessive daytime sleep might be more prevalent in 22q11DS [37-39]. Moreover, Obstructive Sleep Apnea (OSA) might affect up to 40% of individuals with 22q11DS, due to a combination of risk factors including retrognathia, hypotonia and velopharyngeal abnormalities [36, 40-42]. The high rates of both sleep disorders and psychosis make 22q11DS a unique model to investigate the relationship between these two phenomena. Still, the precise sleep phenotype of 22q11DS remains poorly characterized. Most notably, no longitudinal study has associated subjective and objective sleep measures with subsequent vulnerability to psychosis.

A second major challenge of sleep research is that, from a methodological perspective, most studies have independently associated individual subjective or objective sleep variables with a series of clinical variables of interest [43]. Such uni-variate approaches have significant intrinsic limitations in their characterization of both sleep and mental health. Indeed, sleep is a multidimensional phenomenon, implying that individual sleep variables cannot be interpreted in isolation [43]. For instance, a similar reduction in sleep efficiency can be observed in opposite diagnoses of insomnia [44] or narcolepsy [45], according to whether total-sleep-time is reduced or increased. Moreover, sleep disorders are diagnosed from a combination of objective alterations and subjective complaints of unsatisfactory sleep [46], that are neither interchangeable nor univocally correlated [47]. Finally, as pertains to mental health, the effects of sleep disturbances cannot be fully understood unless reciprocal interactions between different psychiatric symptoms are also considered [1]. To address these challenges, we propose to employ two dedicated multivariate approaches, known as Partial-Least-Square-Correlation (PLSC) [48] and Multi-Layer-Network-Analysis (MLNA) [8], which are ideally suited to shed light on the complex pathways linking sleep disturbances to psychosis vulnerability.

We firstly aimed to precisely characterize the sleep phenotype in 22q11DS. Secondly, we aimed to identify multivariate patterns of sleep disturbances associated with symptoms of psychosis. We hypothesized that using a multivariate combination of objective and subjective sleep variables, we would detect meaningful sleep-patterns that would more tightly correlate with psychiatric symptoms. Thirdly, we investigated the impact of sleep disturbances on longitudinal clinical trajectories using recently developed MLNA. We hypothesized that sleep disturbances might specifically influence the affective pathways towards psychosis and might hence provide additional prognostic information when combined to current clinical assessments.

## 2. Methods

### 2.1 Participants

Both healthy controls (HCs) and individuals with 22q11DS were recruited in the context of an ongoing longitudinal study that has been described extensively in previous publications [8, 49]. Participants with 22q11DS were recruited through patient associations or word-of-mouth throughout French and English-speaking European countries. HCs were mostly recruited among non-affected siblings (31/38 HCs). Participants were then followed up longitudinally approximately once every 3 years. All participants and parents provided written informed consent under protocols approved by Ethics Committee Geneva.

For the present study, we included 107 participants, including 69 with 22q11DS (Mean age: 17.5±7.43, age-range: [5.39-34.04], M/F=34:35) and 38 HCs (Mean age: 14.3±6.23 Age-Range: [6.85-31.9], M/F=18:20). For all participants, sleep measures were available at baseline assessment. When exploring clinical correlates of sleep disturbances, we restricted the sample to 46 individuals with 22q11DS for whom the Structured Interview for Psychosis-Risk Syndromes (SIPS) [50] was performed at baseline assessment. In a final longitudinal analysis, we considered a sub-sample of 32 individuals with 22q11DS for whom a longitudinal clinical assessment was available, on average 3.58±0.85 years after baseline assessment. See Table 1 for a full description of demographic features of the sample.

**Table 1:**
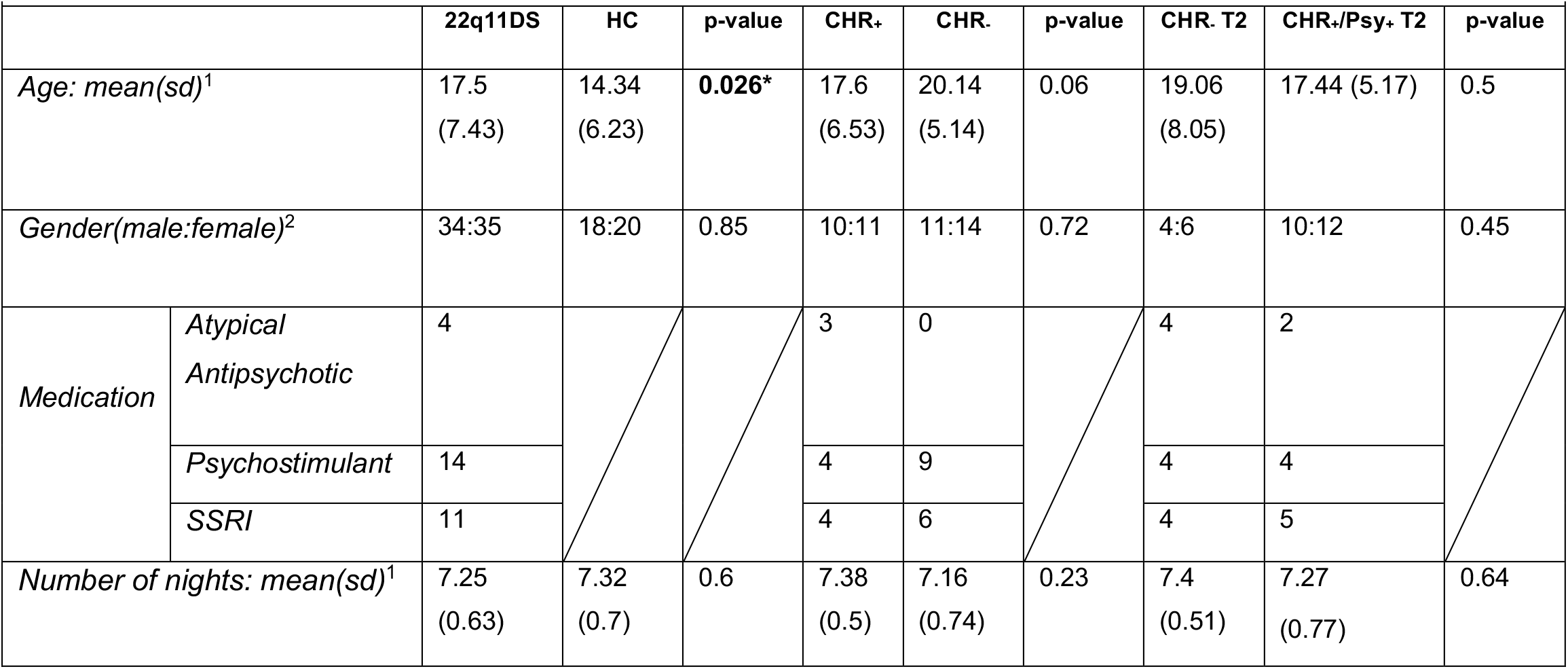
sample characteristics. Demographic information, medication usage at the moment of the visit and number of recorded nights per individual. Differences in age and number of recorded nights per individuals were tested using a two-samples t-test and gender differences were tested using a Chi-squared test. Age differed significantly between 22q11DS and HC. No significant difference in any other groups or characteristics. ^1^ Two-samples t-test ^2^ Chi-squared test * Significant difference (p<0.05)

### 2.2 Sleep measures

Objective sleep measures were collected in a home setting for an average of 7.27±0.65 days using a wrist-worn actigraph (model wGT3X, Actigraph, USA), during the week preceding the clinical assessment. We excluded 2 nights (one for each group) that were shorter than 150 minutes.

Participants and their families also filled in a sleep diary indicating bedtimes and rising times, any nap or removal of the watch. Actigraphy data was visually inspected and subsequently analyzed using the Actilife software (version 6.11.9). Bedtimes and rising times were defined using the sleep diary or, when these were unavailable (77/786 nights), we used the +Sleep autoscoring feature from Actilife. A total of 9 commonly used actigraphy measures were derived from the raw data: sleep-latency, sleep-efficiency, time-spent-in-bed, total-sleep-time, wake-after-sleep-onset (WASO), number-of-awakenings, average-awakening-length, in-bed-time and out-bed-time, using the Sadeh algorithm [51] or the Cole-Kripke algorithm [52], depending on the age of the participant.

Subjective sleep quality during the actigraphy assessment week was measured using the short form of the Children’s Sleep Habits Questionnaire [53]. The questionnaire is subdivided into 4 subparts: bedtime, sleep behavior, morning wake up and waking during the night. Items are rated according to their observed frequency on a 1-to-5 scale. To guarantee consistency across our wide age-range, we adapted the questionnaire for adults and adolescents, by directly addressing the questions to the participants, and removing 8 items that were no longer considered age-appropriate. This yielded 15 items administered across the entire sample.

### 2.3 Clinical measure

Clinical vulnerability to psychosis was assessed using the Structured Interview for Psychosis-risk Syndromes (SIPS) [50], administered by a single trained psychiatrist (SE). The SIPS yields 19 individual items across 4 symptom domains (positive, disorganized, negative and general symptoms), rated on a 1-6 scale according to their severity. To maximize clinical resolution, symptoms were initially considered individually. The SIPS also defines diagnostic criteria that discretely separate individuals considered either non-at-risk of developing psychosis (CHR-negative), at clinical high-risk of developing psychosis (CHR-positive) or fully psychotic (PSY-positive), based on the positive symptom ratings. These discrete criteria were used for confirmatory analyses to define discrete sub-groups, described in Table 1, according to clinical status at baseline (CHR-negative-baseline vs CHR-positive-baseline) and according to clinical status at longitudinal at follow-up (CHR-negative-follow-up vs CHR-positive/PSY-positive-follow-up).

### 2.4 Statistical analysis

All analyses were performed using MATLAB (version R2018a). Prior to all analyses, we corrected for the effects of age and psychotropic medication using linear regression.

#### Univariate analyses

We firstly performed uni-variate comparisons of sleep measures between HCs and 22q11DS using Mixed-Model-Analysis, implemented through a previously published toolbox: https://github.com/danizoeller/myMixedModelsTrajectories [49, 54]. We considered fixed effect of day-of-the-week, diagnosis and day-of-the-week-by-diagnosis interaction and a random intercept per subject. Results were corrected for multiple comparison using the false discovery rate (FDR) [55].

#### Multivariate Partial-Least-Square-Correlation analysis

We then explored the existence of multivariate sleep pattern associated with clinical outcomes, using a publicly available Partial-Least-Square-Correlation (PLSC) algorithm (https://github.com/MIPLabCH/myPLS). PLSC analysis is designed to detect complex multivariate relationships between different data types, such as complex neuroimaging patterns associated with clinical outcomes. Here, all 9 actigraphy measures were firstly averaged across nights and subsequently combined with all 15 subjective sleep measures in a single data-matrix. Sleep variables were then collectively associated with 3 different clinical outcomes: diagnosis of 22q11DS; severity of all 19 SIPS symptoms at baseline; and longitudinal clinical evolution, characterized by combining all 19 SIPS scores at baseline with 19 SIPS scores at longitudinal follow-up. Through singular value decomposition, PLSC extracts sleep patterns, which are associated with clinical outcomes. Statistical significance is established through permutation testing and corrected for multiple comparisons using FDR [56]. Significant patterns are represented as bar-plots, where the contribution (loading) of each variable is represented as bar-height, and bar-direction (up vs down), reflects whether that contribution is positive or negative. Variables that consistently contributed to a specific pattern are identified through bootstrapping and are highlighted in yellow. Finally, each sleep patterns can be condensed into a single sleep score measured in each individual, through matrix multiplication of observed sleep variables with their respective loading. The sleep-score hence represents the extent to which the specific sleep pattern was present in each individual. For confirmatory analysis, we compared such sleep-scores according to discrete clinical cut-offs, using non-parametric permutation testing [57].

#### Multi-Layer-Network-Analysis of Longitudinal Clinical Pathways

We used a recently developed Multi-Layer-Network-Analysis approach to precisely characterize clinical pathways of interaction between symptoms across time [8]. We constructed a single Multi-Layer-Network, computing Spearman correlations between all symptoms, both cross-sectionally at each assessment and longitudinally between baseline and follow-up. Symptoms at each assessment and the sleep-score derived from longitudinal PLSC were then represented as individual network nodes. The spatial position of each node was determined using network dimensionality reduction, whereby Euclidian distance between nodes provides an inverse representation of correlation strength between symptoms, both cross-sectionally and longitudinally. We employed Floyd-Warshall algorithm, implemented in the brain-connectivity toolbox for MATLAB http://www.brain-connectivity-toolbox.net, to compute shortest pathways connecting baseline to follow-up symptoms, across time. We identified variables that mediated a higher proportion of longitudinal clinical pathways than expected by chance and that can hence be considered to disproportionately affect clinical trajectory. By doing so, we evaluated the additive predictive value of baseline sleep disturbances, when combined with all other clinical variables. Moreover, we obtained a precise characterization of clinical pathways mediating the effects of sleep disturbances on vulnerability to psychosis.

## 3. Results

### 3.1 Univariate comparison of sleep variables

Compared to HCs, individuals with 22q11DS presented a significant increase in time-spent-in-bed (Chi^2^=12.71, DF=1, p=0.003), had an earlier in-bed-times (Chi^2^=7.98, DF=2, p=0.047) and longer total-sleep-time (Chi^2^=10.97, DF=1, p=0.004), irrespectively of the day of the week. See figure 1A-C. We also observed a significant non-linear interaction between out-bed-time, diagnosis and the days-of-the-week (Chi^2^=11.59, DF=3, p-interaction-effect=0.01) leading individuals with 22q11DS to present a significant delay in rising times, particularly on Saturday mornings (Chi^2^=11.59, DF=4, p-group-effect=0.047). See figure 1D.

**Figure 1:**
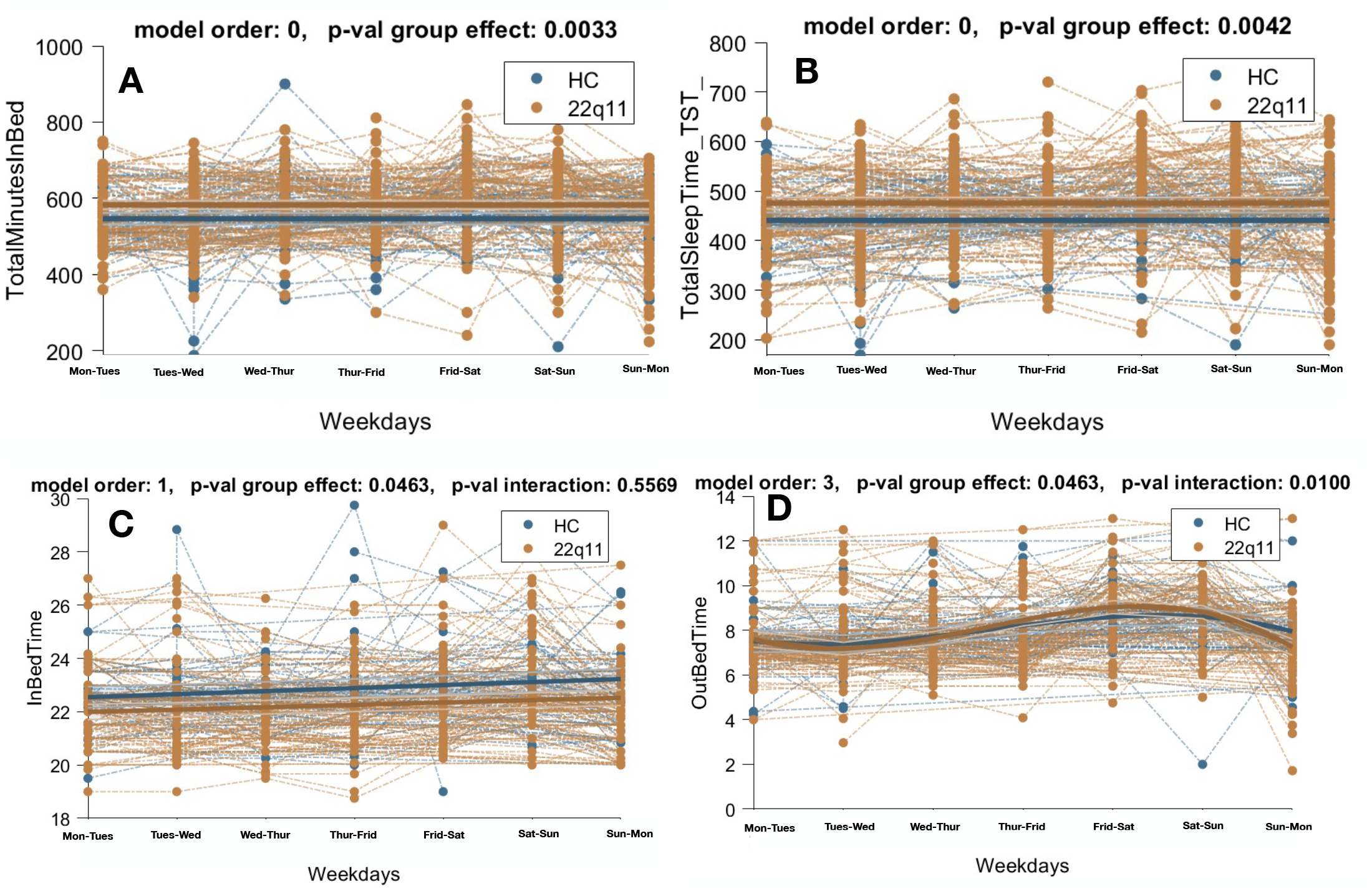
Univariate comparison of objective sleep variables between Healthy Controls (HCs) and individuals with 22q11DS, as function of week-days using Mixed Models Linear Regression. **Panel A:** Differences in Total Minutes in Bed across samples. **Panel B:** Differences in Total Sleep Time across samples. **Panel C:** Differences in In-Bed-Time across samples. **Panel D:** Differences in Out-Bed-Time across samples.

### 3.2 Multivariate sleep pattern associated with 22q11DS

PLSC identified a single sleep pattern that significantly differentiated individuals with 22q11DS from HCs (p=0.002, R=0.55) and was characterized by earlier in-bed-time and longer time-spent-in-bed combined with reduced number-of-awakenings, leading to longer total-sleep-time. See figure 2. Interestingly, despite longer sleep duration, and subjective reports of more regular sleep schedule, the 22q11DS sleep pattern was associated with higher levels of subjective daytime fatigue. This sleep pattern was also characterized by more frequent fear the dark and teeth-grinding.

**Figure 2:**
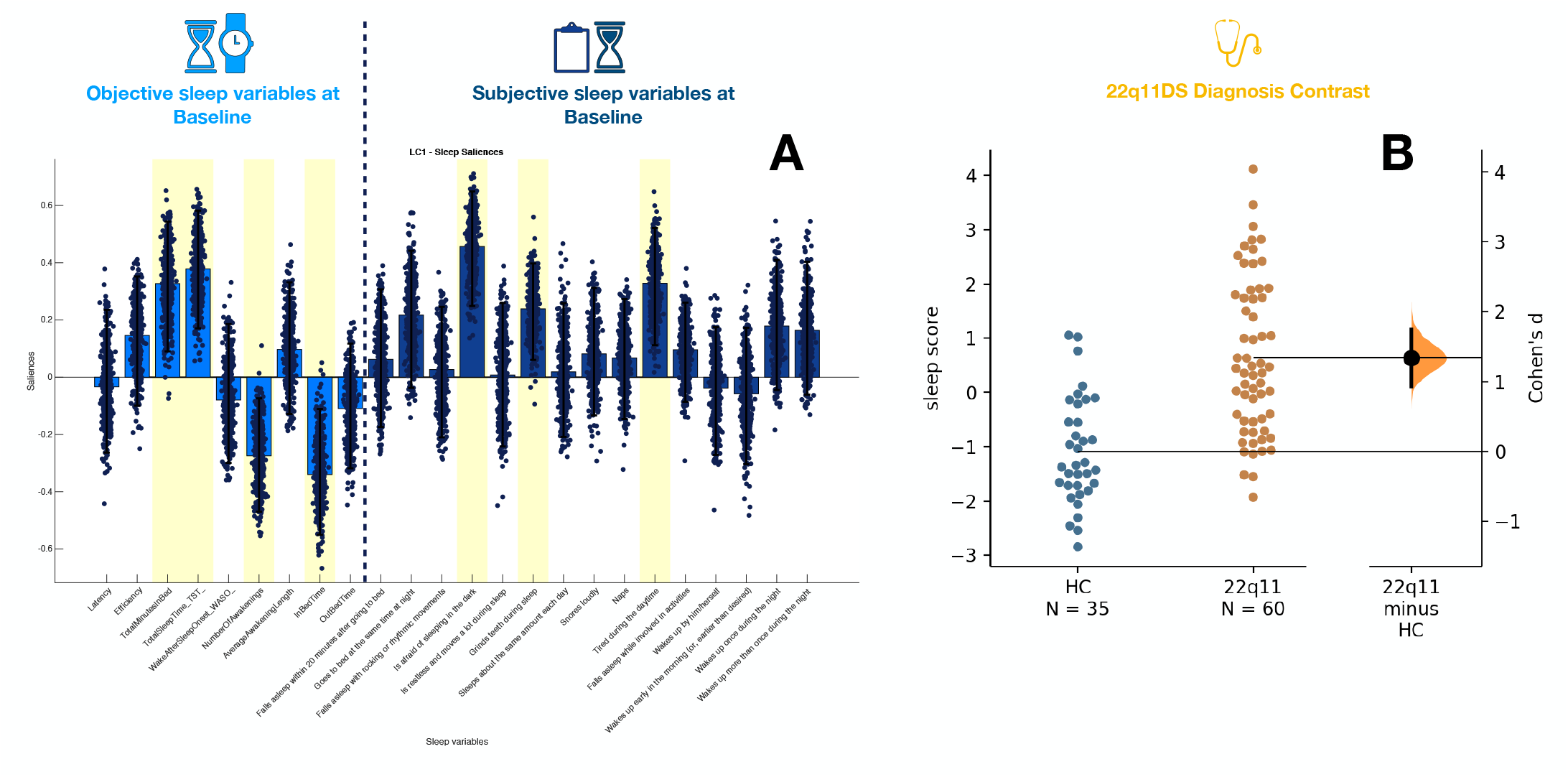
Multi-Variate PLSC analysis of sleep patterns associated with 22q11DS. **Panel A:** Sleep pattern composed of objective and subjective sleep variables associated with 22q11DS. The loading of each variable, capturing its contribution to the sleep pattern, is represented as the height of the bar plot. Objective sleep variables, derived from actigraphy are plotted on the left in light blue. Subjective sleep variables, derived from questionnaires, are plotted on the right in dark blue. The direction of the bar (up vs down) reflects the positive vs negative contribute of the specific variable to sleep pattern. Hence, a positive loading reflects a tendency of the variable to be increased in 22q11DS compared to HCs, whereas a negative loading reflects an opposite reduction. Scatter plots represent the distribution of the loading of a specific variable according to 500 iterations of bootstrapping of the original sample. Variables highlighted in yellow are considered to have a stable contribution to the sleep pattern, as captured by a consistent positive or negative loading, throughout à 95% confidence interval of the distribution of bootstrapped loadings. **Panel B:** Differences in sleep-scores across HCs and 22q11DS. Each point represents the sleep score computed in each subject capturing the extent to which the sleep pattern represented in Panel A was present in each individual.

### 3.3 Multivariate sleep pattern associated with clinical symptoms at baseline

PLSC analysis identified a single significant sleep pattern that correlated strongly with the intensity of SIPS symptoms at baseline (p=0.001, R=0.80). See Figure 3C. The sleep pattern loaded strongly on subjective measures of daytime somnolence and fatigue, despite objectively reduced sleep-latency and increased total-sleep-time. Symptoms commonly reported in the context of OSA, including loud snoring, multiple awakenings and agitated sleep also strongly contributed to the pattern. Subjective reports of falling asleep difficulties and fear of the dark also contributed, albeit less strongly. See Figure 3A.

**Figure 3:**
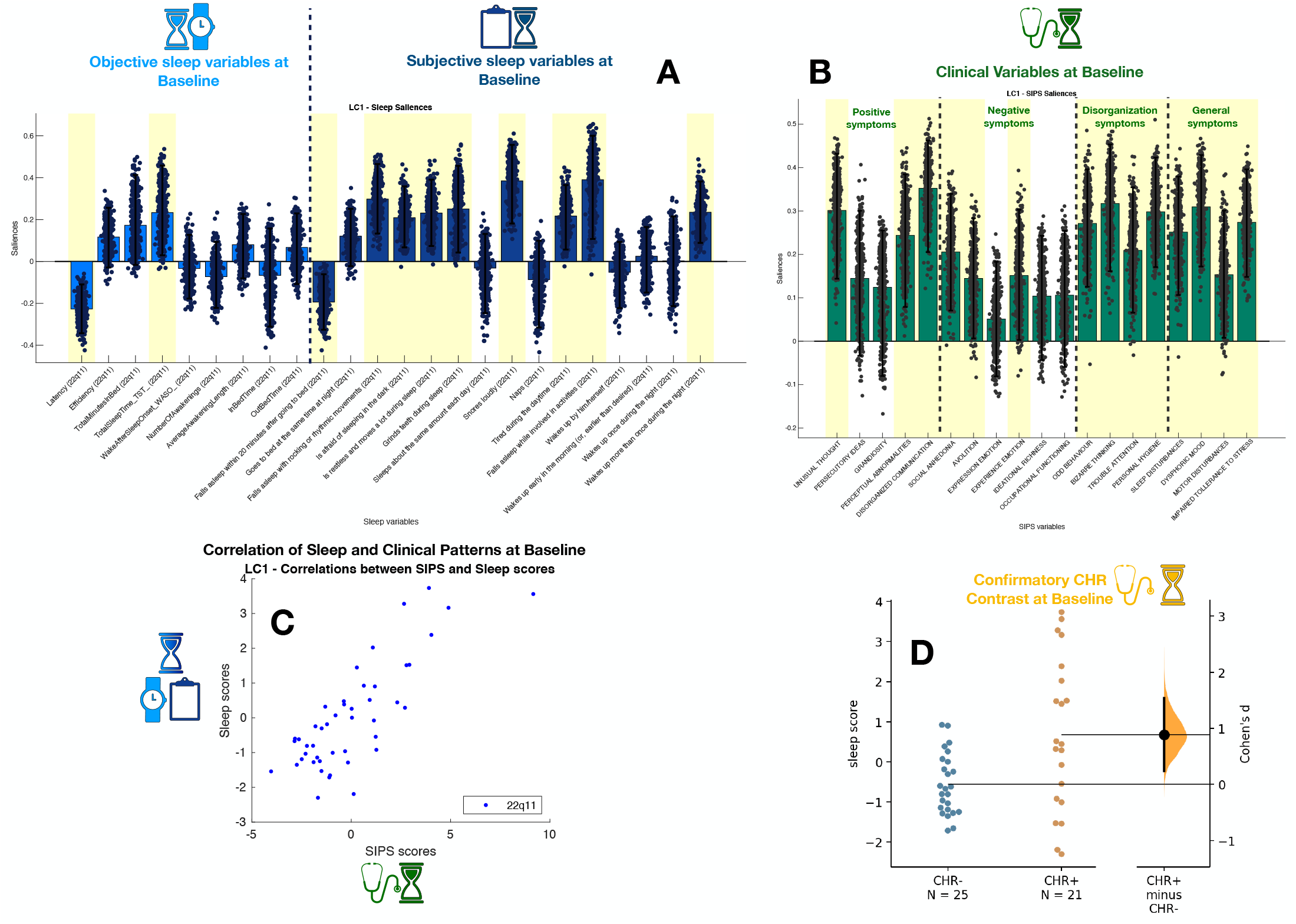
Multi-Variate PLSC analysis of sleep patterns associated with SIPS symptoms of vulnerability to psychosis at baseline assessment. **Panel A:** Sleep pattern composed of objective and subjective sleep variables associated with SIPS symptoms at baseline. The loading of each variable, capturing its contribution to the sleep pattern, is represented as the height of the bar plot. Objective sleep variables, derived from actigraphy are plotted on the left in light blue. Subjective sleep variables, derived from questionnaires, are plotted on the right in dark blue. The direction of the bar (up vs down) reflects the positive vs negative contribute of the specific variable to sleep pattern. Hence, a positive loading reflects a tendency of the variable to be positively correlated to symptom intensity, whereas a negative loading reflects an opposite reduction. Scatter plots represent the distribution of the loading of a specific variable according to 500 iterations of bootstrapping of the original sample. **Panel B:** Clinical pattern representing the contribution of each SIPS symptom at baseline to the association with sleep variables. As in Panel A, variables highlighted in yellow are considered to have a stable contribution to the sleep pattern, as captured by a coherent positive or negative contribution, throughout à 95% confidence interval of the bootstrapped loadings. **Panel C:** Correlation of sleep scores and clinical scores across subjects. Sleep scores and clinical scores reflect the extent to which the sleep or clinical pattern was represented in each individual. The correlation of sleep scores and clinical scores reflects the strength of the association between multivariate sleep and clinical patterns. **Panel D:** Differences in sleep-scores between individuals with 22q11DS separated according to discrete CHR criteria at baseline.

The clinical pattern that correlated with disrupted sleep was characterized by a broad combination of several core psychotic and disorganization symptoms, including unusual-thought-content, disorganized-communication, bizarre-thinking and odd-behavior, as well general symptoms such as dysphoric-mood and impaired-tolerance-to-stress. See Figure 3B. A Confirmatory analysis revealed that the corresponding sleep-score was strongly increased in CHR-positive compared to non-CHR individuals (p<0.0001, Cohen’s D: 0.864). See Figure 3D.

### 3.4 Multivariate sleep pattern associated with longitudinal clinical trajectories

Longitudinal PLSC identified a single component that strongly explained the association between baseline sleep variables and clinical trajectory, measured with a combination of baseline and follow-up symptoms (p=0.001, R=0.83) See Figure 4C.

**Figure 4:**
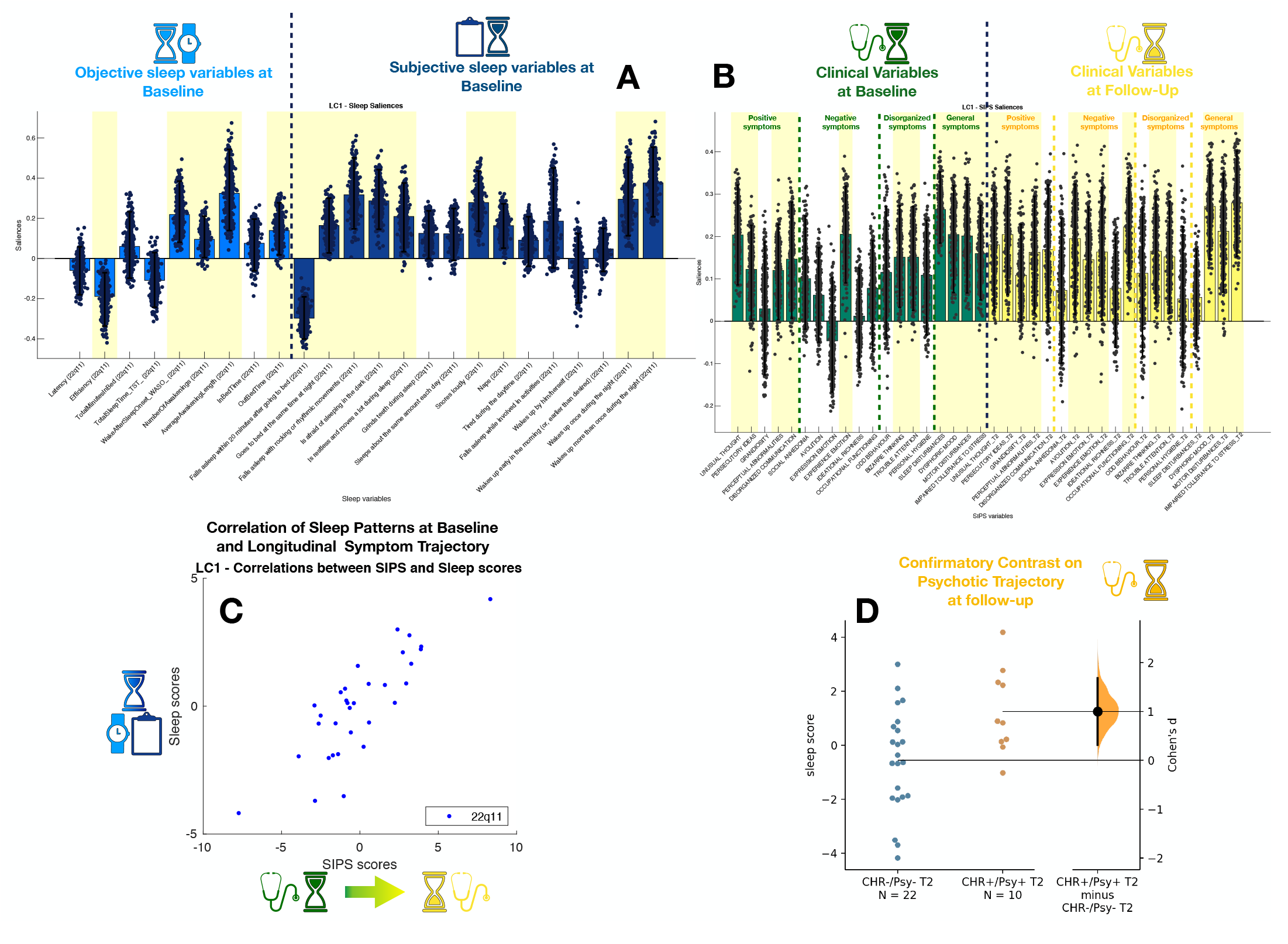
Multi-Variate PLSC analysis of sleep patterns associated with longitudinal clinical trajectory of SIPS symptoms from baseline to follow-up assessments **Panel A:** Sleep pattern composed of objective and subjective sleep variables associated with SIPS symptoms at baseline and follow-up. The loading of each variable, capturing its contribution to the sleep pattern, is represented as the height of the bar plot. Objective sleep variables, derived from actigraphy are plotted on the left in light blue. Subjective sleep variables, derived from questionnaires, are plotted on the right in dark blue. The direction of the bar (up vs down) reflects the positive vs negative contribute of the specific variable to sleep pattern. Hence, a positive loading reflects a tendency of the variable to be positively correlated to symptom intensity, whereas a negative loading reflects an opposite reduction. Scatter plots represent the distribution of the loading of a specific variable according to 500 iterations of bootstrapping of the original sample. **Panel B:** Clinical pattern representing the contribution of each SIPS symptom at baseline and follow-up to the association with sleep variables. Symptoms at baseline assessment are represented in green on the left part of the graph. Symptoms at follow-up assessment are represented in yellow on the right part of the graph. As in Panel A, variables highlighted in yellow are considered to have a stable contribution to the sleep pattern, as captured by a coherent positive or negative contribution, throughout à 95% confidence interval of the bootstrapped loadings. **Panel C:** Correlation of sleep scores and longitudinal clinical scores across subjects. Sleep scores and clinical scores reflect the extent to which the sleep or clinical pattern was represented in each individual. The correlation of sleep scores and clinical scores reflects the strength of the association between multivariate sleep and clinical patterns. **Panel D:** Differences in sleep-scores between individuals with 22q11DS separated according to longitudinal clinical trajectory at follow-up assessment.

The sleep pattern loaded strongly on subjective markers of increased daytime somnolence combined with symptoms commonly reported in the context of OSA, including loud snoring, agitated sleep, and more frequent subjective awakenings. This was associated with an objective increase in average-awakening-length and wake-after-sleep-onset, combined with reduced sleep-efficiency and total-sleep-time, suggestive of increased sleep fragmentation. See Figure 4A.

At baseline, such sleep pattern was associated with some positive symptoms, including unusual-thought-content, and most general symptoms. Interestingly, clinical associations became broader and stronger at longitudinal follow-up, with significant loading on most positive and negative symptoms and a strong loading of the general symptoms of dysphoric-mood and impaired-tolerance-to-stress See Figure 4B. The disrupted-sleep-score was also significantly increased among individuals who were either CHR-positive or fully psychotic at longitudinal follow-up, compared to individuals who were either consistently non-CHR or for whom CHR-criteria had remitted (p<0.0001, Cohen’s d: 0.849). See Figure 4D.

We then used Multi-Layer-Network-Analysis to dissect pathways linking sleep disturbances to longitudinal clinical trajectory. Symptoms at both assessments are represented as network nodes and plotted according to network dimensionality reduction. Hence, a shorted Euclidian distances between network nodes reflects stronger correlation between corresponding symptoms (R=-0.43, p<0.0001). The main horizontal dimension separated positive and general symptoms, located on the left side of the graph, from negative and disorganization symptoms located on the right side. See Figure 5. The second vertical dimension, mainly describes the longitudinal time dimensions, separating baseline symptoms on the bottom of the graph from follow-up symptoms, located on the upper part of the graph. The overall structure of the network was extremely consistent with what previously described [8].

**Figure 5:**
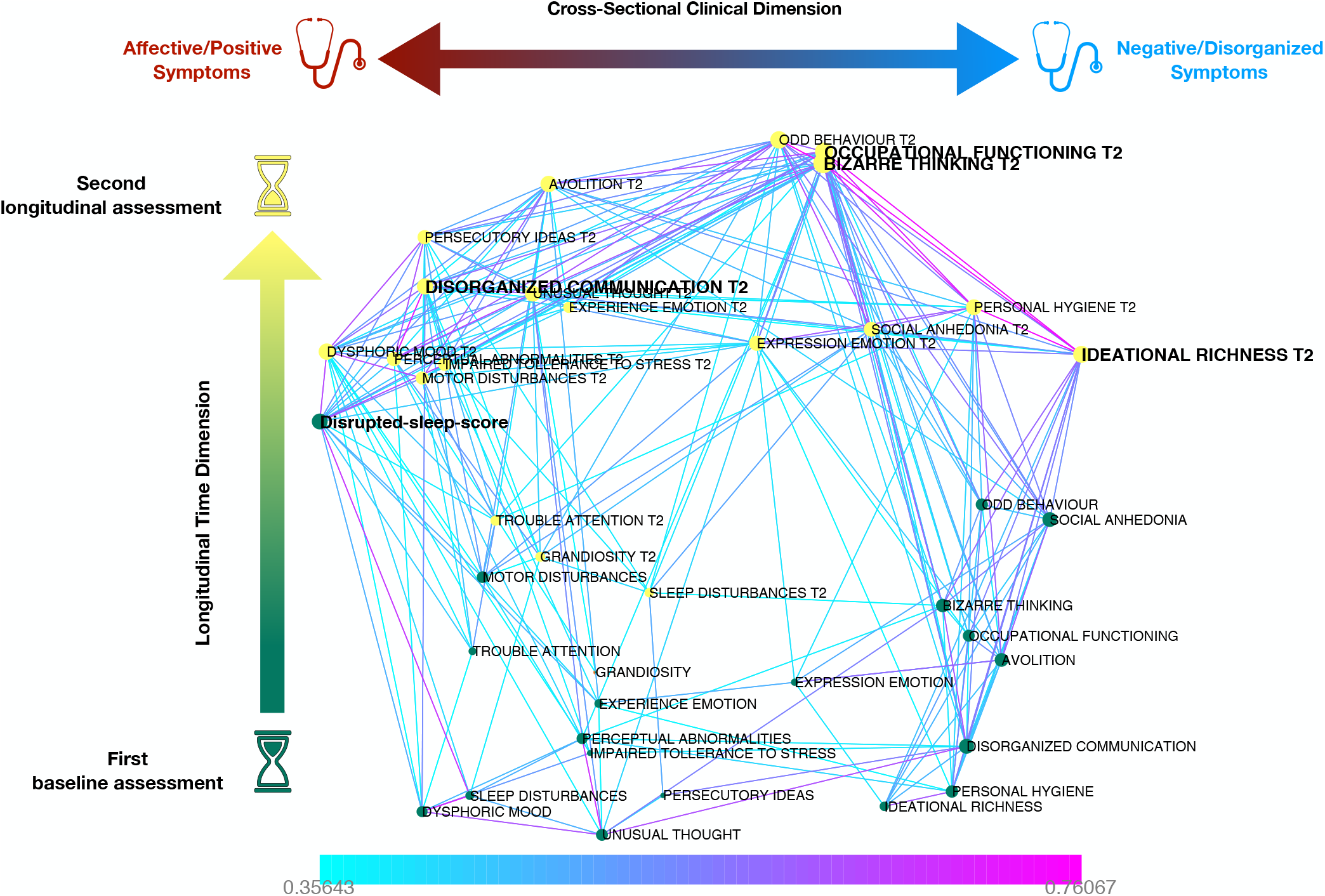
Multilayer Temporal Network Analysis capturing both cross-sectional and longitudinal associations between SIPS symptoms at baseline and follow-up assessments. The sleep score derived from longitudinal PLSC analysis is considered as an additional network node. Variables measured at baseline are plotted as individual network nodes in green. Symptoms at longitudinal follow-up are plotted in yellow. The spatial position of each variable is derived from network dimensionality reduction. The first network dimension plotted along the horizontal axis separates affective and positive symptoms located on the left from negative and disorganized symptoms, located on the right. The second network dimension separates symptoms at baseline, located on the bottom of the graph from symptoms at longitudinal follow-up located on the upper part of the graph. Nodes that are highlighted in bold are significant longitudinal network hubs, that mediate a disproportionate number of clinical pathways linking baseline to follow-up symptoms, across time.

The disrupted-sleep-score was located on the left side of the graph, reflecting strong correlations with positive and affective symptoms. Interestingly, the disrupted-sleep-score occupied an intermediary position along the vertical temporal dimension, located closer to follow-up than to baseline symptoms, in the upper part of network. This suggests that sleep disturbances are more tightly associated with clinical trajectory at longitudinal follow-up, than to concomitant clinical difficulties estimated at baseline.

Remarkably, graph-theory revealed that the disrupted-sleep-score was the only baseline variable that mediated a disproportionate number of clinical pathways across time (p=0.002). This suggests that the disrupted-sleep-score might provide additional prognostic value, even when considering all other clinical variables measuring vulnerability to psychosis. In accordance with previous reports [8], the algorithm also identified several follow-up symptoms that acted as funnels for previous psychopathology, including disorganized communication (p=0.003), bizarre-thinking (p<0.001), reduced-occupational-functioning (p<0.001) and reduced-ideational-richness (p<0.001).

We then represented only clinical paths that traversed through the disrupted-sleep-score. See Figure 6. Sleep disturbances mediated the effects of baseline attention and affective difficulties including dysphoric-mood and reduced-experience-of-emotion, on the persistence of homologous affective symptoms at follow-up. Interestingly, sleep disturbances also mediated the effects of affective symptoms on the subsequent development of broader difficulties, including hallucinations or unusual-thought-content, and core negative symptoms such as avolition and reduced-occupational-functioning. As such, disrupted sleep could be characterized as a gateway towards subsequent psychopathology.

**Figure 6:**
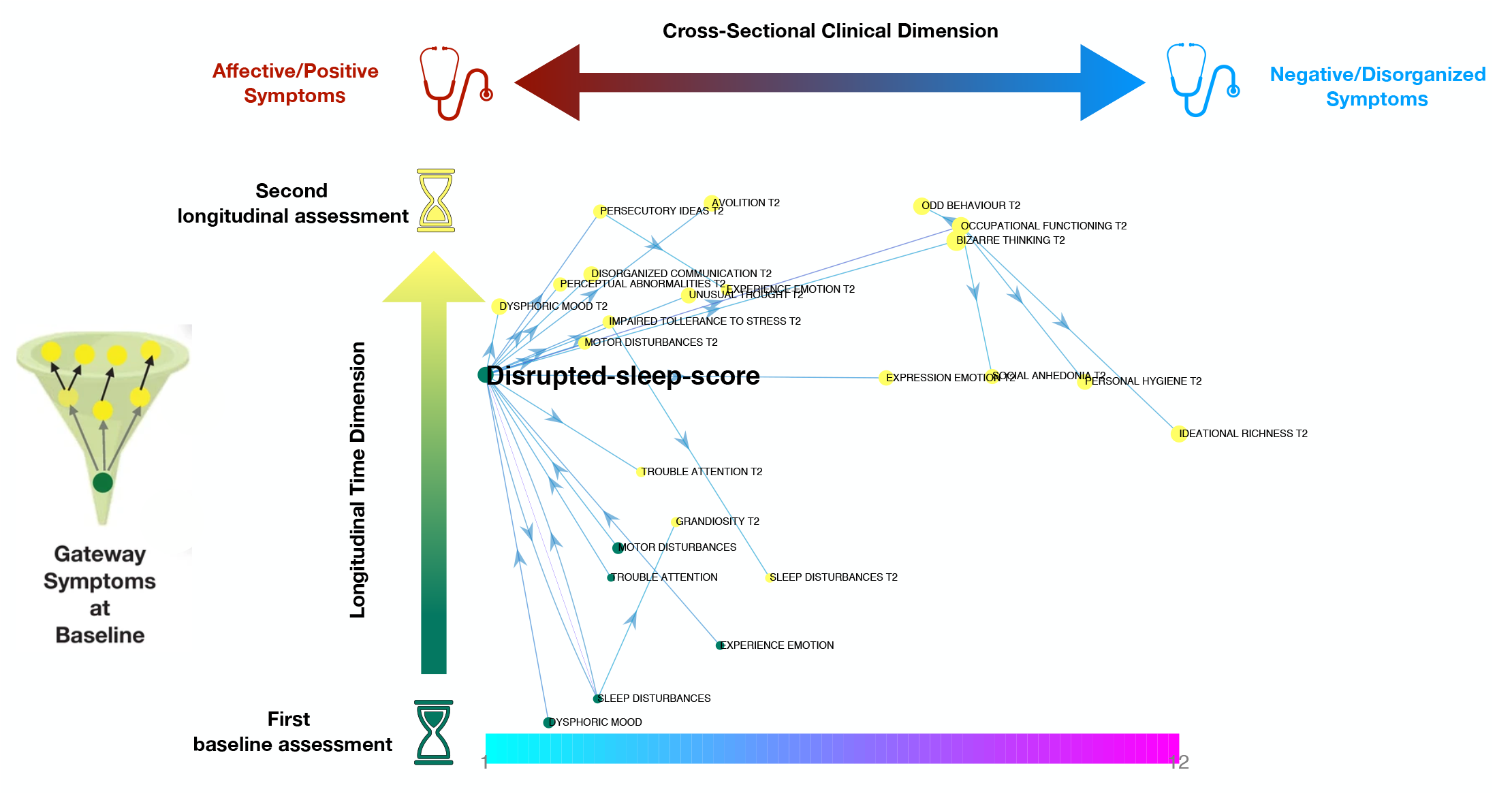
Multilayer Temporal Network Analysis representing longitudinal clinical pathways between symptoms that are mediated by the Disrupted-sleep-score, measured at baseline. The sleep score captures the extent to which the specific sleep pattern, evocative of OSA, was present in each individual with 22q11DS. A high sleep score mediated the effects of baseline symptoms of affective disturbance, represented in green in the bottom left part of the graph on persistence of such affective symptoms at longitudinal follow-up, represented in the upper left part of the graph, and the subsequent development of positive and negative/disorganised symptoms, located in the upper central and right part of the graph.

## 4. Discussion

In the present study, we conducted for the first time a comprehensive characterization of sleep quality in 22q11DS, using both subjective and objective sleep measures and novel multivariate approaches. We firstly delineated a specific sleep phenotype suggestive of non-restorative sleep in 22q11DS. Secondly, we described a disrupted-sleep-pattern that was broadly associated with the severity of psychotic symptoms and overall psychopathology. Thirdly, we showed that disrupted sleep strongly predicts longitudinal clinical trajectory towards psychosis in 22q11DS. Finally, we dissected clinical pathways linking sleep disturbances to psychotic vulnerability, using a dedicated network analysis approach. These results suggest that sleep quality represents a novel promising therapeutic target that could significantly influence clinical trajectory in 22q11DS.

Our results indicate that individuals with 22q11DS present a specific sleep-pattern, characterized mainly by increased daytime somnolence despite objectively longer sleep duration. These results could indicate that sleep is insufficiently restorative in 22q11DS, resulting in increased sleep pressure and requiring compensation strategies, such as earlier bedtimes and delayed rising-times. Insufficiently restorative sleep can be observed in primary hypersomnia, or in relation to intrinsic sleep disorders affecting sleep architecture. Actigraphy does not allow to reliably ascertain sleep architecture. However, the only previous polysomnography study in 22q11DS also observed an increased total-sleep-time associated with altered sleep architecture, including increased REM sleep latency, and an increase in N1 sleep [58]. Moreover, 22q11DS might affect sleep architecture due to risk of OSA, which has been observed in up to 40% of individuals with the syndrome [38, 40, 42, 59]. Remarkably, the clinical impact of OSA on the psychiatric and neurocognitive phenotype of 22q11DS remains largely unknown.

When associating sleep variables with psychiatric symptoms, we again observed a pattern of daytime somnolence despite reduced sleep-latency and increased sleep-time, suggestive of non-restorative sleep and increased sleep pressure. Interestingly, non-restorative-sleep was associated with symptoms that are classically reported in the context of OSA, including loud snoring and subjective impression of agitated and disrupted sleep. Remarkably, this disrupted-sleep-pattern was broadly associated with more severe psychotic symptoms, together with symptoms of affective psychopathology. The disrupted sleep pattern was moreover strongly overrepresented among CHR individuals. These results could suggest that disrupted sleep evocative of OSA, might contribute to psychiatric difficulties observed in 22q11DS, including both affective and psychotic symptoms.

Recently, there has been a renowned interest in the relationship between OSA and psychosis [60]. A large epidemiological study reported an association between OSA and increased risk of both mood and anxiety disorders as well as a 35% increase in risk of psychosis [61]. The existing screening studies, performed in relatively small samples, suggest that the prevalence of OSA among psychotic patients is between 12.5% and 25%, which is twice as frequent than what is observed in HCs [21, 62, 63]. Until now, the association between OSA and psychosis has been mostly interpreted in relation to shared risk factors, including mainly increased BMI [62, 64]. Some authors have suggested that symptoms of OSA, including daytime sleepiness, might mimic negative and cognitive symptoms of schizophrenia [65]. However, no longitudinal study has evaluated whether the presence of OSA might increase vulnerability to psychosis among at-risk individuals.

22q11DS is associated with increased risk of both OSA and psychosis, making it a unique model to explore the longitudinal relationship between the two disorders. Our longitudinal analysis identified a sleep pattern that was strongly evocative of OSA, including excessive daytime fatigue, snoring and subjective and objective markers of agitated sleep. Remarkably, this sleep pattern strongly predicted longitudinal persistence or worsening of psychotic manifestations. We used a recently developed Multi-Layer-Network-Analysis to precisely characterize longitudinal pathways linking disrupted sleep to psychosis vulnerability [8]. Firstly, this analysis revealed that sleep disruption was a key mediator influencing clinical trajectories, even when considering all other clinical variables. This would suggest that sleep phenotyping might provide additional prognostic value, even when combined to current gold-standard assessments of psychosis vulnerability. Interestingly, the disrupted-sleep-score specifically mediated the relationship between baseline symptoms of emotional dysregulation, both on the subsequent persistence of emotional symptoms and the subsequent development of psychotic and negative symptoms. These results suggest that OSA might actively influence the clinical trajectory, in particular by promoting an affective pathway towards psychosis [7].

Current models of such affective pathway propose that predisposition to negative affect, observed in early stages of psychosis, results from a state of amygdala hyperactivation, related to altered top-down pre-frontal connectivity and aggravated by environmental stress [66]. Prolonged amygdala hyperactivity could then lead to sub-cortical-dopaminergic and hippocampal hyperactivity observed in psychosis, either by direct influencing GABAergic transmission [67-69] or through indirect effects of hypercortisolemia [70]. This neurobiological pathway has been recently observed in 22q11DS, where anxiety is related to amygdala hyperactivity and amygdala-prefrontal dysconnectivity [71]. Heightened negative affect predicted psychotic vulnerability in 22q11DS, both through amygdala-striatal hyperconnectivity [72] and through altered HPA-axis and hippocampal maturation [33]. Interestingly, as reviewed above, a wealth of studies have demonstrated that sleep disruption, in particular affecting REM sleep, can determine emotional dysregulation through a combination of amygdala and sub-cortical-dopaminergic hyperactivity and HPA-axis/noradrenergic dysregulation [12]. As such, our results could suggest that presence of OSA, which particularly affects REM sleep [73], could accentuate limbic and dopaminergic hyperactivity, predisposing affected individuals to persistent emotional dysregulation and hence increasing risk of progression along the affective pathway towards psychosis. Moreover, genetically-driven alterations in limbic GABA-ergic transmission, observed in animal models of 22q11DS [74], could make individuals with the syndrome particularly vulnerable to the effects of sleep disturbances. These hypotheses could be empirically tested with longitudinal studies combining sleep and clinical phenotyping with neuroimaging, as well as in animal models of the syndrome.

If confirmed, these results carry significant implications for clinical practice and future research. Firstly, they provide a strong rational for systematic screening of OSA and overall sleep quality in 22q11DS, as it is currently recommended for other neurogenetic conditions [75]. Indeed, OSA can be successfully treated in both neurogenetic conditions [76, 77] and idiopathic psychosis [78]. Future research should evaluate the feasibility and clinical impact of treating OSA in 22q11DS. Indeed, our results would suggest that management of sleep disturbances could significantly affect long term psychiatric outcome in the syndrome. Our results also support the need for a more systematic investigation of the relationship between disrupted sleep, including OSA, and vulnerability to psychosis to inform clinical practice. Finally, from a methodological perspective, our results suggest that a multivariate longitudinal approach, that coherently integrates multiple sleep variables, could help shed light on complex pathways linking sleep disruption to developmental psychopathology.

Our results should be interpreted considering several limitations. Firstly gold-standard polysomnography was not available. Consequently, we could measure neither altered sleep architecture nor presence of OSA, rendering our interpretations concerning mechanism underlying the disrupted-sleep-patterns necessarily speculative. Secondly, sleep measures were not available at the longitudinal follow-up, not allowing us to measure the impact of previous psychopathology on subsequent sleep disturbance. Finally, we cannot formally exclude that additional factors might have contributed to our results. We did however make considerable efforts, detailed as supplementary analyses, to explore the contributions of the frequently reported confounds, including age, BMI and psychotropic medications.

## Data Availability

All data produced in the present study are available upon reasonable request to the authors

## Acknowledgments

This study was supported by the Swiss National Science Foundation (SNSF) (Grant numbers: to SE FNS 320030_179404, FNS 324730_144260) and by the National Center of Competence in Research (NCCR) Synapsy-The Synaptic Bases of Mental Diseases (SNF, Grant number: 51AU40_125759). MarS (#163859), MauS (#162006) and CorrS (#209096) were supported by grants from the SNF. We warmly thank all the families who participated in the study.

## References

1. Freeman, D., et al., Sleep disturbance and psychiatric disorders. The Lancet Psychiatry, 2020. 7(7): p. 628–637.

2. Borsboom, D., A network theory of mental disorders. World Psychiatry, 2017. 16(1): p. 5–13.

3. Caspi, A., et al., The p Factor: One General Psychopathology Factor in the Structure of Psychiatric Disorders? Clin Psychol Sci, 2014. 2(2): p. 119–137.

4. Kendell, R. and A. Jablensky, Distinguishing between the validity and utility of psychiatric diagnoses. Am J Psychiatry, 2003. 160(1): p. 4–12.

5. McGorry, P. and B. Nelson, Why We Need a Transdiagnostic Staging Approach to Emerging Psychopathology, Early Diagnosis, and Treatment. JAMA Psychiatry, 2016. 73(3): p. 191–2.

6. Hartmann, J.A., et al., At-risk studies and clinical antecedents of psychosis, bipolar disorder and depression: a scoping review in the context of clinical staging. Psychol Med, 2019. 49(2): p. 177–189.

7. Myin-Germeys, I. and J. van Os, Stress-reactivity in psychosis: evidence for an affective pathway to psychosis. Clin Psychol Rev, 2007. 27(4): p. 409–24.

8. Sandini, C., et al., Characterization and prediction of clinical pathways of vulnerability to psychosis through graph signal processing. Elife, 2021. 10.

9. van Os, J., The dynamics of subthreshold psychopathology: implications for diagnosis and treatment. Am J Psychiatry, 2013. 170(7): p. 695–8.

10. Braun, U., A Network Perspective on the Search for Common Transdiagnostic Brain Mechanisms. Biol Psychiatry, 2018. 84(6): p. e47–e48.

11. Carragher, N., et al., Disorders without borders: current and future directions in the meta-structure of mental disorders. Soc Psychiatry Psychiatr Epidemiol, 2015. 50(3): p. 339–50.

12. Goldstein, A.N. and M.P. Walker, The role of sleep in emotional brain function. Annu Rev Clin Psychol, 2014. 10: p. 679–708.

13. Palmer, C.A. and C.A. Alfano, Sleep and emotion regulation: An organizing, integrative review. Sleep Med Rev, 2017. 31: p. 6–16.

14. Spruyt, K., A review of developmental consequences of poor sleep in childhood. Sleep Med, 2019. 60: p. 3–12.

15. Hegerl, U. and T. Hensch, The vigilance regulation model of affective disorders and ADHD. Neurosci Biobehav Rev, 2014. 44: p. 45–57.

16. Pandi-Perumal, S.R., et al., Clarifying the role of sleep in depression: A narrative review. Psychiatry Res, 2020. 291: p. 113239.

17. Watling, J., et al., Sleep Loss and Affective Functioning: More Than Just Mood. Behav Sleep Med, 2017. 15(5): p. 394–409.

18. Kaskie, R.E. and F. Ferrarelli, Sleep disturbances in schizophrenia: what we know, what still needs to be done. Curr Opin Psychol, 2020. 34: p. 68–71.

19. Faulkner, S. and P. Bee, Experiences, perspectives and priorities of people with schizophrenia spectrum disorders regarding sleep disturbance and its treatment: a qualitative study. BMC Psychiatry, 2017. 17(1): p. 158.

20. Ong, W.J., et al., Association between sleep quality and domains of quality of life amongst patients with first episode psychosis. Health and Quality of Life Outcomes, 2020. 18(1).

21. Reeve, S., B. Sheaves, and D. Freeman, Sleep Disorders in Early Psychosis: Incidence, Severity, and Association With Clinical Symptoms. Schizophr Bull, 2019. 45(2): p. 287–295.

22. Chemerinski, E., et al., Insomnia as a predictor for symptom worsening following antipsychotic withdrawal in schizophrenia. Compr Psychiatry, 2002. 43(5): p. 393–6.

23. Lunsford-Avery, J.R., D.J. Dean, and V.A. Mittal, Self-reported sleep disturbances associated with procedural learning impairment in adolescents at ultra-high risk for psychosis. Schizophr Res, 2017. 190: p. 160–163.

24. Lunsford-Avery, J.R., et al., Adolescents at clinical-high risk for psychosis: Circadian rhythm disturbances predict worsened prognosis at 1-year follow-up. Schizophr Res, 2017. 189: p. 37–42.

25. Lunsford-Avery, J.R., et al., Actigraphic-measured sleep disturbance predicts increased positive symptoms in adolescents at ultra high-risk for psychosis: A longitudinal study. Schizophr Res, 2015. 164(1-3): p. 15–20.

26. Reeve, S., et al., Sleep duration and psychotic experiences in patients at risk of psychosis: A secondary analysis of the EDIE-2 trial. Schizophr Res, 2019. 204: p. 326–333.

27. Ferrarelli, F., Sleep Abnormalities in Schizophrenia: State of the Art and Next Steps. Am J Psychiatry, 2021. 178(10): p. 903–913.

28. Insel, T.R., Rethinking schizophrenia. Nature, 2010. 468(7321): p. 187–93.

29. Blagojevic, C., et al., Estimate of the contemporary live-birth prevalence of recurrent 22q11.2 deletions: a cross-sectional analysis from population-based newborn screening. CMAJ Open, 2021. 9(3): p. E802–E809.

30. Robin, N.H. and R.J. Shprintzen, Defining the clinical spectrum of deletion 22q11.2. J Pediatr, 2005. 147(1): p. 90–6.

31. Schneider, M., et al., Psychiatric disorders from childhood to adulthood in 22q11.2 deletion syndrome: results from the International Consortium on Brain and Behavior in 22q11.2 Deletion Syndrome. Am J Psychiatry, 2014. 171(6): p. 627–39.

32. Armando, M., et al., Coping Strategies Mediate the Effect of Stressful Life Events on Schizotypal Traits and Psychotic Symptoms in 22q11.2 Deletion Syndrome. Schizophr Bull, 2018. 44(Suppl_2): p. S525–S535.

33. Sandini, C., et al., Pituitary dysmaturation affects psychopathology and neurodevelopment in 22q11.2 Deletion Syndrome. Psychoneuroendocrinology, 2020. 113: p. 104540.

34. Gothelf, D., et al., Risk factors for the emergence of psychotic disorders in adolescents with 22q11.2 deletion syndrome. Am J Psychiatry, 2007. 164(4): p. 663–9.

35. Gothelf, D., et al., Risk factors and the evolution of psychosis in 22q11.2 deletion syndrome: a longitudinal 2-site study. J Am Acad Child Adolesc Psychiatry, 2013. 52(11): p. 1192-1203.e3.

36. Arganbright, J.M., et al., Sleep patterns and problems among children with 22q11 deletion syndrome. Mol Genet Genomic Med, 2020. 8(6): p. e1153.

37. Hyde, J., et al., Gene Deletion and Sleep Depletion: Exploring the Relationship Between Sleep and Affect in 22q11.2 Deletion Syndrome. J Genet Psychol, 2021. 182(5): p. 304–316.

38. Moulding, H.A., et al., Sleep problems and associations with psychopathology and cognition in young people with 22q11.2 deletion syndrome (22q11.2DS). Psychol Med, 2020. 50(7): p. 1191–1202.

39. O’Hora, K.P., et al., Copy number variation at the 22q11.2 locus influences prevalence, severity, and psychiatric impact of sleep disturbance. J Neurodev Disord, 2022. 14(1): p. 41.

40. Kennedy, W.P., et al., 22q11.2 Deletion syndrome and obstructive sleep apnea. Int J Pediatr Otorhinolaryngol, 2014. 78(8): p. 1360–4.

41. Lee, A., et al., Defining Risk of Postoperative Obstructive Sleep Apnea in Patients With 22q11.2DS Undergoing Pharyngeal Flap Surgery for Velopharyngeal Dysfunction Using Polysomnographic Evaluation. Cleft Palate Craniofac J, 2020. 57(7): p. 808–818.

42. Silvestre, J., et al., Screening for obstructive sleep apnea in children with syndromic cleft lip and/or palate. J Plast Reconstr Aesthet Surg, 2014. 67(11): p. 1475–80.

43. van de Langenberg, S.C.N., D. Kocevska, and A.I. Luik, The multidimensionality of sleep in population-based samples: a narrative review. J Sleep Res, 2022. 31(4): p. e13608.

44. Baglioni, C., et al., Sleep changes in the disorder of insomnia: a meta-analysis of polysomnographic studies. Sleep Med Rev, 2014. 18(3): p. 195–213.

45. Zhang, Y., et al., Polysomnographic nighttime features of narcolepsy: A systematic review and meta-analysis. Sleep Med Rev, 2021. 58: p. 101488.

46. Sateia, M.J., International classification of sleep disorders-third edition: highlights and modifications. Chest, 2014. 146(5): p. 1387–1394.

47. Cudney, L.E., et al., Investigating the relationship between objective measures of sleep and self-report sleep quality in healthy adults: a review. J Clin Sleep Med, 2022. 18(3): p. 927–936.

48. Krishnan, A., et al., Partial Least Squares (PLS) methods for neuroimaging: a tutorial and review. Neuroimage, 2011. 56(2): p. 455–75.

49. Mancini, V., et al., Positive psychotic symptoms are associated with divergent developmental trajectories of hippocampal volume during late adolescence in patients with 22q11DS. Mol Psychiatry, 2019.

50. Miller, T.J., et al., Prospective diagnosis of the initial prodrome for schizophrenia based on the Structured Interview for Prodromal Syndromes: preliminary evidence of interrater reliability and predictive validity. Am J Psychiatry, 2002. 159(5): p. 863–5.

51. Sadeh, A., K.M. Sharkey, and M.A. Carskadon, Activity-based sleep-wake identification: an empirical test of methodological issues. Sleep, 1994. 17(3): p. 201–7.

52. Cole, R.J., et al., Automatic sleep/wake identification from wrist activity. Sleep, 1992. 15(5): p. 461–9.

53. Bonuck, K.A., et al., Modified Children’s sleep habits questionnaire for behavioral sleep problems: A validation study. Sleep Health, 2017. 3(3): p. 136–141.

54. Mutlu, A.K., et al., Sex differences in thickness, and folding developments throughout the cortex. NeuroImage, 2013. 82: p. 200–207.

55. Benjamini, Y. and D. Yekutieli, False Discovery Rate–Adjusted Multiple Confidence Intervals for Selected Parameters. Journal of the American Statistical Association, 2005. 100(469): p. 71–81.

56. Zoller, D., et al., Disentangling resting-state BOLD variability and PCC functional connectivity in 22q11.2 deletion syndrome. Neuroimage, 2017. 149: p. 85–97.

57. Ho, J., et al., Moving beyond P values: data analysis with estimation graphics. Nat Methods, 2019. 16(7): p. 565–566.

58. Mauro, J., et al., Analysis of REM sleep without atonia in 22q11.2 deletion syndrome determined by domiciliary polysomnography: a cross sectional study. Sleep, 2022. 45(2).

59. Crockett, D.J., et al., Obstructive sleep apnea syndrome in children with 22q11.2 deletion syndrome after operative intervention for velopharyngeal insufficiency. Front Pediatr, 2014. 2: p. 84.

60. Myles, H., et al., Obstructive sleep apnea and schizophrenia: A systematic review to inform clinical practice. Schizophr Res, 2016. 170(1): p. 222–5.

61. Sharafkhaneh, A., et al., Association of psychiatric disorders and sleep apnea in a large cohort. Sleep, 2005. 28(11): p. 1405–11.

62. Myles, H., et al., Obstructive sleep apnoea is more prevalent in men with schizophrenia compared to general population controls: results of a matched cohort study. Australas Psychiatry, 2018. 26(6): p. 600–603.

63. Lane, A.R., et al., Screening for obstructive sleep apnoea in an early psychosis cohort: a pilot study. Australas Psychiatry, 2020. 28(2): p. 180–185.

64. Liu, D., et al., Risk Factors for Obstructive Sleep Apnea Are Prevalent in People with Psychosis and Correlate with Impaired Social Functioning and Poor Physical Health. Front Psychiatry, 2016. 7: p. 139.

65. Lane, A.R., S. Foley, and D. Siskind, We snooze, they lose: where is the conversation about obstructive sleep apnoea in early psychosis? Australas Psychiatry, 2019. 27(3): p. 314.

66. Taylor, S.F., et al., The Fragile Brain: Stress Vulnerability, Negative Affect and GABAergic Neurocircuits in Psychosis. Schizophr Bull, 2019. 45(6): p. 1170–1183.

67. Akirav, I. and G. Richter-Levin, Biphasic modulation of hippocampal plasticity by behavioral stress and basolateral amygdala stimulation in the rat. J Neurosci, 1999. 19(23): p. 10530–5.

68. Benes, F.M., et al., Regulation of the GABA cell phenotype in hippocampus of schizophrenics and bipolars. Proc Natl Acad Sci U S A, 2007. 104(24): p. 10164–9.

69. Berretta, S., D.W. Munno, and F.M. Benes, Amygdalar activation alters the hippocampal GABA system: “partial” modelling for postmortem changes in schizophrenia. J Comp Neurol, 2001. 431(2): p. 129–38.

70. Walker, E., V. Mittal, and K. Tessner, Stress and the hypothalamic pituitary adrenal axis in the developmental course of schizophrenia. Annu Rev Clin Psychol, 2008. 4: p. 189–216.

71. Zoller, D., et al., Large-Scale Brain Network Dynamics Provide a Measure of Psychosis and Anxiety in 22q11.2 Deletion Syndrome. Biol Psychiatry Cogn Neurosci Neuroimaging, 2019.

72. Delavari, F., et al., 2022.

73. Spruyt, K. and D. Gozal, REM and NREM sleep-state distribution of respiratory events in habitually snoring school-aged community children. Sleep Med, 2012. 13(2): p. 178–84.

74. Meechan, D.W., et al., Modeling a model: Mouse genetics, 22q11.2 Deletion Syndrome, and disorders of cortical circuit development. Prog Neurobiol, 2015. 130: p. 1–28.

75. Knollman, P.D., et al., Adherence to Guidelines for Screening Polysomnography in Children with Down Syndrome. Otolaryngol Head Neck Surg, 2019. 161(1): p. 157–163.

76. Maris, M., et al., Outcome of adenotonsillectomy in children with Down syndrome and obstructive sleep apnoea. Arch Dis Child, 2017. 102(4): p. 331–336.

77. Simpson, R., et al., Obstructive sleep apnea in patients with Down syndrome: current perspectives. Nat Sci Sleep, 2018. 10: p. 287–293.

78. Giles, J.J., et al., Obstructive Sleep Apnea Is Treatable With Continuous Positive Airway Pressure in People With Schizophrenia and Other Psychotic Disorders. Schizophr Bull, 2022. 48(2): p. 437–446.

