## Supplementary material for "Multivariate patterns of disrupted sleep longitudinally predict affective vulnerability to psychosis in 22q11.2 Deletion Syndrome"

Supplementary analysis 1: PLSC analyses of sleep patterns associated with age

**Methods**

As a first supplementary analysis, using the partial-least-square-correlation (PLSC) (see statistical analysis in methods for more details), we contrasted sleep variables against age, diagnosis, and age by diagnosis interaction, correcting for the effect of psychotropic medication with linear regression.

**Results**

*PLSC analysis of sleep pattern associated with age*

This analysis detected a first component that explained the most the co-variance between sleep and behavioural variables (p=0.001, R=0.69). Specifically, The PLSC detected a strong effect of age on both subjective and objective sleep parameters. See supplementary figure 1B.

Age exerted a negative effect on objective measures of sleep latency, sleep duration, awakenings as well as the subjective measure of being afraid of the dark. Variables which on the other hand increased with age included objective measures of sleep efficiency, in-bed-times and subjective measures of falling asleep with rhythmic movement and naps. These results confirm that age is the main factor contributing to changes in subjective and objective sleep parameters, in accordance with previous literature [1]. The effect of age was not significantly different between HCs and 22q11DS as captured by a non-significant loading on the age-by-diagnosis variable. These results justify the importance of accounting for the effect of age with linear regression as done in the main text, in order to detect clinical correlates of sleep disturbance. See supplementary figure 1A.

The second component showed a strong association between objective and subjective sleep measures with 22q11DS as well as a weaker association with age-by-diagnosis-interaction (p=0.006, R=0.55). See supplementary figure 2B. The sleep pattern strongly resembled what described in main-text as differentiating 22q11DS from HCs, loading on the objective measure of increased sleep duration and reduced number of awakenings as well as subjective measure of being more afraid of sleeping in the dark, teeth-grinding, feeling tired during the day and increased awakenings. A weak age-by-diagnosis contrast could mean that differences among samples might already be present at a young age and become subsequently less evident. However, given that our HCs and 22q11DS were not precisely matched for age, with fewer adult HCs, we are not confident in drawing strong conclusions from this weak age-by-diagnosis interaction effect.

**Conclusion**

Overall, these results show confirm that age has a strong effect on objective and subjective sleep parameters, which is mostly similar across HCs and 22q11DS individuals. Accounting for the effect of age is hence important for maximise the detection of clinical correlates of sleep disturbances. Moreover, our results show that the sleep pattern that differentiates 22q11DS from HCs is already present from an early age.

Supplementary analysis 1: PLSC analyses of sleep patterns associated with medication

**Methods**

As a second supplementary analysis, using the PLSC, we contrasted sleep variables against the 3 main types of psychotropic medication, namely atypical antipsychotics, Selective Serotine Reuptake Inhibitors (SSRIs) and psychostimulant medication. This analysis was conducted only in the 22q11DS sample, as no psychotropic medication had been prescribed to HCs, and corrected for the effect of age with linear regression.

*PLSC analysis of sleep pattern associated with medication*

The PLSC revealed the existence of two components accounting for the co-variation of mediation use and sleep parameters.

The first component loaded only on use of antipsychotics (p=0.005, R=0.69). The use of antipsychotics was associated with increased sleep duration and longer awakenings, reduced number of awakenings and earlier bedtimes as well as subjective measures of increased sleep regularity, snoring and daytime somnolence.

We also detected a second component describing an association between sleep variables and use of both psychostimulant and SSRI (p=0.016, R=0.54). The sleep pattern loaded only on objective sleep variables, including decreased efficiency and sleep duration as well as increased awakenings’ length.

**Discussion**

The sleep pattern associated with the use of antipsychotic medication resembled the pattern described in the main text as being associated with psychotic symptoms, and evocative of OSA. This result confirms the importance of accounting for the effects of antipsychotic medications, as was performed in the main text. However, the fact that clinical correlates of sleep disruption remain strongly significant even after accounting for use of antipsychotic medication suggests that medication is not solely responsible for sleep disruption. These results could suggest that correlation between use of antipsychotics and sleep disruption could be partially accounted for by the higher propensity of more symptomatic patients to be medicated. An intriguing complementary hypothesis is that the prolonged use antipsychotic medication might accentuate preexisting sleep disturbances and contribute to sub-optimal clinical outcome.

A second interesting finding is that both psychostimulant and SSRI exerted a differential effect on objective sleep parameters, being associated with an opposite reduction of total-sleep-time and in-bed-time, compared to antipsychotics. This is differential pattern is expected, given the wake-promoting effects that characterize both psychostimulants and SSRI medications [2]. On the other hand, a tendency to be associated with longer average-awakening-length is shared across both sleep patterns. Interestingly, however, the objective sleep alterations associated with psychostimulant or SSRI medication are not associated with subjective sleep variables, such as daytime somnolence. This would suggest that sleep alterations associated with psychostimulant and SSRI medication might be better tolerated both subjectively and potentially also in terms of clinical corelates.

**Conclusions:**

These results suggest that different types of psychotropic medication are associated with differential objective and subjective sleep alterations in 22q11DS. They confirm the importance of accounting for the effects of psychotropic medication when interpreting clinical correlates of sleep disruption, as was done in the main text. The fact that clinical correlates of sleep disturbance remain significant when medication was accounted for suggests that use of psychotropic medication is not the sole factor determining sleep disruption in 22q11DS.

Supplementary analysis 3: Association between sleep score and BMI z-score

**Methods**

As a confirmatory analysis, we explored the association between the sleep scores reported by the PLSC and the BMI z-scores of individuals with 22q11DS. The individual sleep scores represent to which extent an individual’s sleep variables correspond to the sleep pattern reported by the PLSC (see figure 3A and 4A for the sleep patterns). Z-scores were used to normalize BMI values across ages. BMI z-scores were calculated using the BMI-for-age charts from the CDC growth standards [3].

**Results**

There was no significant correlation between the sleep scores extracted from the cross-sectional PLSC (between sleep variables at baseline and SIPS variables at baseline) and BMI z-score (Pearson correlation coefficient ρ=0.16, p=0.3) as well as between the sleep score extracted from longitudinal PLSC (between sleep variables at baseline and SIPS variables at baseline and follow-up) and BMI z-score (Pearson correlation coefficient ρ=0.1, p=0.59). See supplementary figure 3A and 3B.

**Discussion**

This analysis was aimed to measure the association between BMI scores and the disrupted sleep evocative of OSA. These results suggest that in 22q11DS, a higher BMI, which might be a consequence of antipsychotic medication, is not the sole contributor to this disrupted sleep patterns. Additional factors including variation in velopharyngeal anatomy might play a more significant role in determining sleep disruption in 22q11DS.

1.	Ohayon, M.M., et al., *Meta-Analysis of Quantitative Sleep Parameters From Childhood to Old Age in Healthy Individuals: Developing Normative Sleep Values Across the Human Lifespan.* Sleep, 2004. **27**(7): p. 1255-1273.

2.	Hegerl, U. and T. Hensch, *The vigilance regulation model of affective disorders and ADHD.* Neurosci Biobehav Rev, 2014. **44**: p. 45-57.

3.	Kuczmarski, R.J., et al., *2000 CDC Growth Charts for the United States: methods and development.* Vital Health Stat 11, 2002(246): p. 1-190.

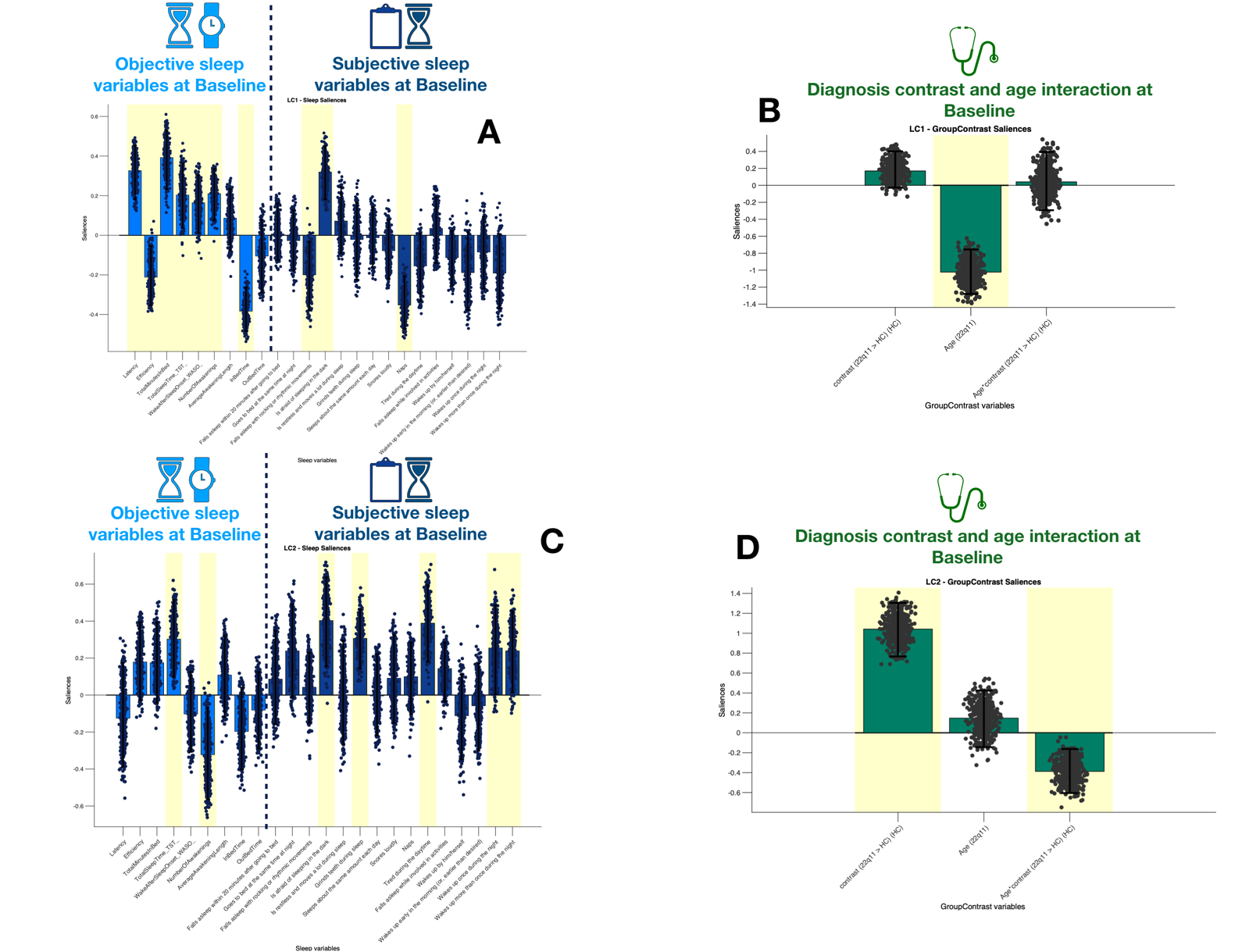

**Supplementary figure 1:** Multi-Variate PLSC analysis of sleep patterns associated with 22q11DS, age and age by diagnosis interaction, two components. **Panel A and C:** Sleep patterns composed of objective and subjective sleep variables. The loading of each variable, capturing its contribution to the sleep pattern, is represented as the height of the bar plot. Objective sleep variables, derived from actigraphy are plotted on the left in light blue. Subjective sleep variables, derived from questionnaires, are plotted on the right in dark blue. The direction of the bar (up vs down) reflects the positive vs negative contribute of the specific variable to sleep pattern. Scatter plots represent the distribution of the loading of a specific variable according to 500 iterations of bootstrapping of the original sample. Variables highlighted in yellow are considered to have a stable contribution to the sleep pattern, as captured by a consistent positive or negative loading, throughout à 95% confidence interval of the distribution of bootstrapped loadings. **Panel A**: First component, a positive loading reflects a tendency of the variable to be negatively correlated with the age, whereas a negative loading reflects an opposite reduction. **Panel C**: Second component, a positive loading reflects a tendency of the variable to be increased in 22q11DS and to decrease with age in 22q11DS, whereas a negative loading reflects an opposite reduction. **Panel B and D:** Clinical pattern representing the contribution of diagnosis, age and diagnosis by age interaction at baseline to the association with sleep variables. As in Panel A and C, variables highlighted in yellow are considered to have a stable contribution to the sleep pattern, as captured by a coherent positive or negative contribution, throughout à 95% confidence interval of the bootstrapped loadings. Panel B is associated with Panel A, representing the first component and panel D with panel C, representing the second component.

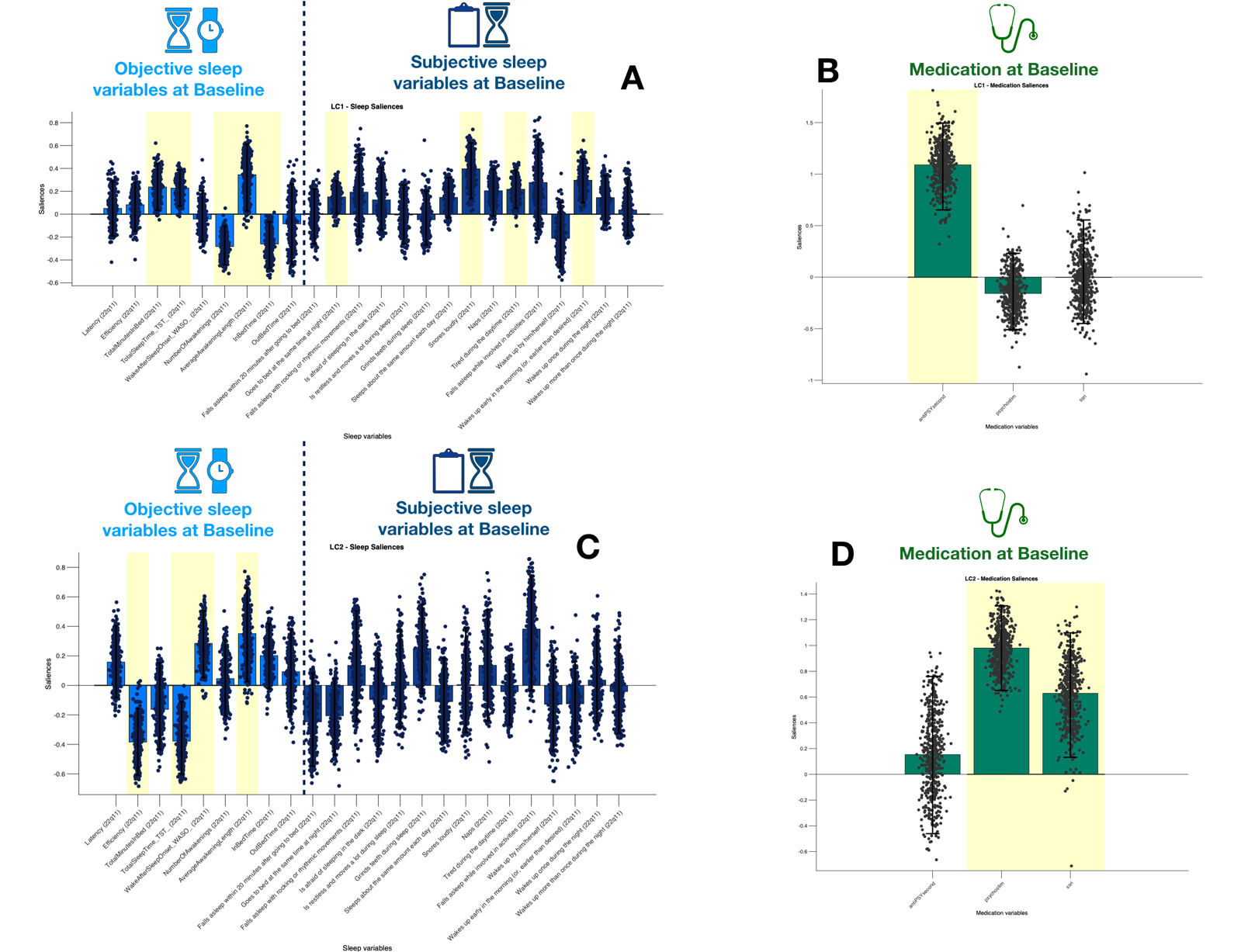
 **Supplementary figure 2:** Multi-Variate PLSC analysis of sleep patterns associated with medication (atypical antipsychotic, psychostimulant and Selective Serotonin Reuptake Inhibitors (SSRI)), two components. **Panel A and C:** Sleep patterns composed of objective and subjective sleep variables associated with the medication. The loading of each variable, capturing its contribution to the sleep pattern, is represented as the height of the bar plot. Objective sleep variables, derived from actigraphy are plotted on the left in light blue. Subjective sleep variables, derived from questionnaires, are plotted on the right in dark blue. The direction of the bar (up vs down) reflects the positive vs negative contribute of the specific variable to sleep pattern. Scatter plots represent the distribution of the loading of a specific variable according to 500 iterations of bootstrapping of the original sample. Variables highlighted in yellow are considered to have a stable contribution to the sleep pattern, as captured by a consistent positive or negative loading, throughout à 95% confidence interval of the distribution of bootstrapped loadings. **Panel A**: First component, a positive loading reflects a positive correlation with atypical antipsychotic, whereas a negative loading reflects an opposite reduction. **Panel C**: Second component, a positive loading reflects a tendency of the variable to be positively correlated with psychostimulants and SSRI, whereas a negative loading reflects an opposite reduction. **Panel B and D:** Medication pattern representing the contribution of the medication at baseline to the association with sleep variables. As in Panel A and C, variables highlighted in yellow are considered to have a stable contribution to the sleep pattern, as captured by a coherent positive or negative contribution, throughout à 95% confidence interval of the bootstrapped loadings. Panel B is associated with Panel A, representing the first component and panel D with panel C, representing the second component.

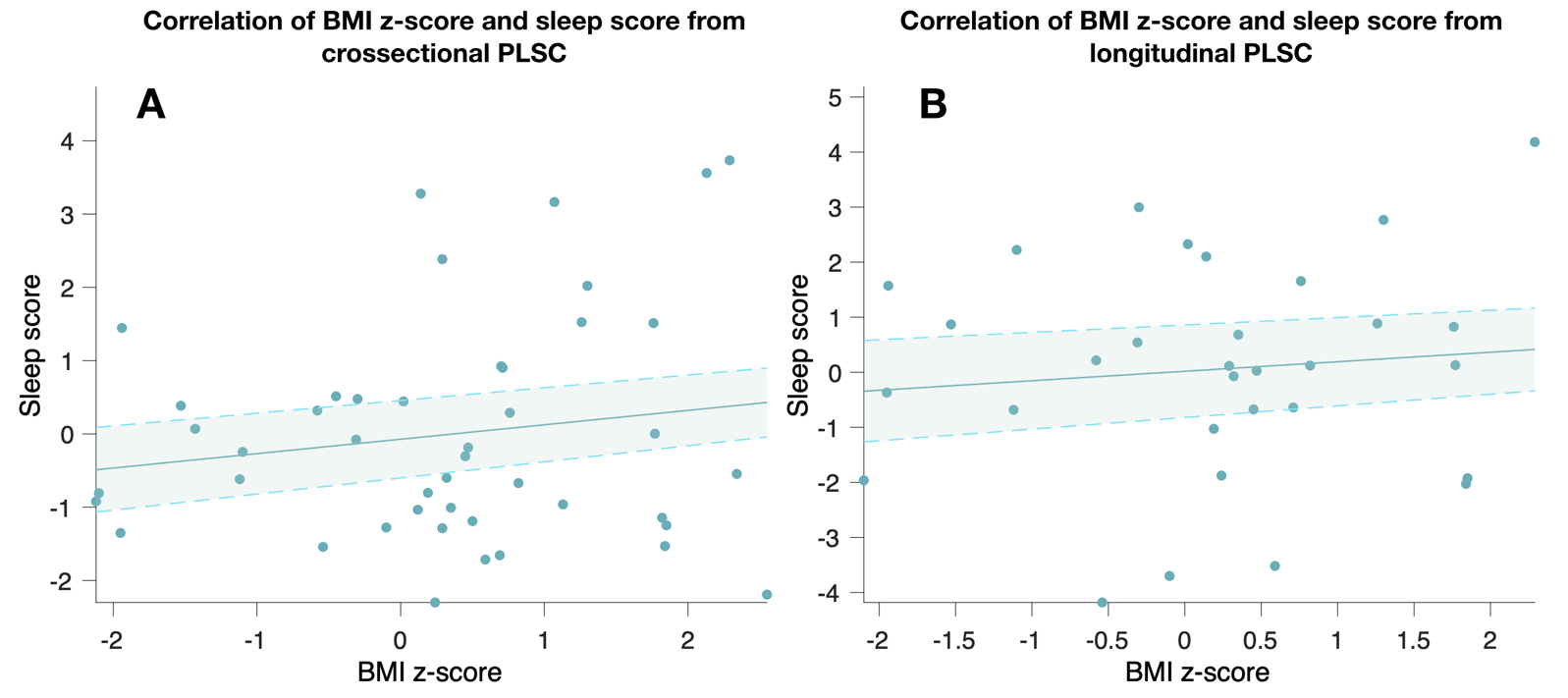

**Supplementary figure 3:** Association between sleep scores and BMI z-scores. **Panel A:** Association between the sleep scores extracted from the crossectional PLSC (between sleep variables at baseline and SIPS variables at baseline) and BMI z-score. Each dot represents an individual affected by 22q11DS. The higher the sleep score, the closer the sleep variables of the individual to the sleep pattern reported by the crossectional PLSC (see figure 3A). **Panel B**: Correlation between sleep score extracted from longitudinal PLSC (between sleep variables at baseline and SIPS variables at baseline and follow-up) and BMI z-score. Each dot represents an individual affected by 22q11DS. The higher the sleep score, the closer the sleep variables of the individual to the sleep pattern reported by the longitudinal PLSC (see figure 4A).
